# Novel Perceval Sizing Technique: Single Center Experience

**DOI:** 10.1101/2024.08.06.24311534

**Authors:** Rafik Margaryan, Giovanni Concistre, Giacomo Bianchi, Marco Solinas

## Abstract

**Objective:** Aim of this study was to compare old (manufacturer recommended) and new (institution based) sizing techniques for sutureless valve.

**Materials:** A 226 consecutively operated patients underwent aortic valve replacement with Perceval sutureless valve (Corcym) and had CT scan with contrast enhancement were included to this study. The final decision of appropriate size is based on intra-operative obturator sizing. Briefly, we have measured on the CT scans the annular ring surface and perimeter in order to estimate the prosthesis size. New sizing technique uses only white obturator of Perceval and it should passe trough the annulus with slight friction, which practically under-sizes with respect to manufacturer’s recommendations.

**Results:** The operative mortality was 1 (0.4424779%). There were no prosthesis migration neither annular rupture in any group. The mean follow up was lower in new group (3.3 ± 2.0 vs 9.7 ± 1.6, p < 0.01). At the discharge the patients who have used the new sizing technique had less gradient on the prosthetic valve (13.4 ± 5.0 vs 15.2 ± 5.5, p = 0.02). The new sizing was less prone to degeneration at the follow-up which would require intervention (13.4 ± 5.0 vs 15.2 ± 5.5, p = 0.02). Oversizing of 22.6% had significant role on valve gradient increase and structural degeneration (*p* < 0.05).

**Conclusions:** New sizing technique is safe and reproducible. It seem to deliver better immediate and long term benefits for Perceval sutureless valve, less postoperative gradient, less probability or structural degeneration.

## Introduction

The Perceval sutureless aortic bioprosthesis (Corcym Inc.) is a intraanular, true sutureless valve implanted surgically. Similarly to transcatheter aortic valve implantation (TAVI) devices, the anchoring and good sealing of the Perceval bioprosthesis relies on oversizing the stent frame compared with the native aortic annulus. Cardiac computed tomography (CT) is a gold standard technique for measuring the aortic annulus in patients undergoing TAVI, and the CT-derived axial image of the aortic virtual basal ring (VBR) is considered as the reference for sizing by most of the manufacturers of transcatheter valves [1]. Interestingly, the VBR lies exactly on the plane passing through the nadir of the 3 aortic cusps, that is where, according to the instructions for use, a correctly positioned Perceval valve should be deployed [2]. Because the Perceval valve stent is a self-expandable nitinol stent with relatively low radial force, the area of the aortic annulus VBR could provide a good estimate of the final cross-sectional area of the stent after deployment and could therefore be used to calculate the degree of oversizing (or underexpansion) of the valve. Recent evidence in Perceval treated patients suggests that significant over-sizing can increased transprosthetic gradients and possibly predisposing to other negative outcomes such as valve thrombosis, low platelet counts, thromboembolic events, and early degeneration [3]. Moreover, there is evidence that excessive oversizing of the Perceval valve is detrimental [4]. However, a systematic analysis of the relationship between the degree of oversizing and the hemodynamic performance and longevity of the Perceval valve is lacking. The present study investigats novel sizing technique in order to find best evidence base valve diameter determination.

## Material and Methods

### Patients

From March 2011 to July 2023, 1150 patients underwent aortic valve replacement (AVR) with the Perceval at our institution [5]. A baseline preoperative cardiac CT allowing for correct sizing of the aortic annulus was available for 226 patients constituted the population of the present study. The demographic and clinical patients’ characteristics are reported in Table 1. All of the data presented in the study were prospectively collected and entered in our institutional database, that are filled in consecutively by anesthesiologists, surgeons, perfusionists, and intensive care unit and ward physicians. The study was approved by the Ospedale Del Cuore Fondazone G Monasterio Clinical Audit Committee to meet ethical and legal requirements, and individual consent was waived.

**Table 1:**
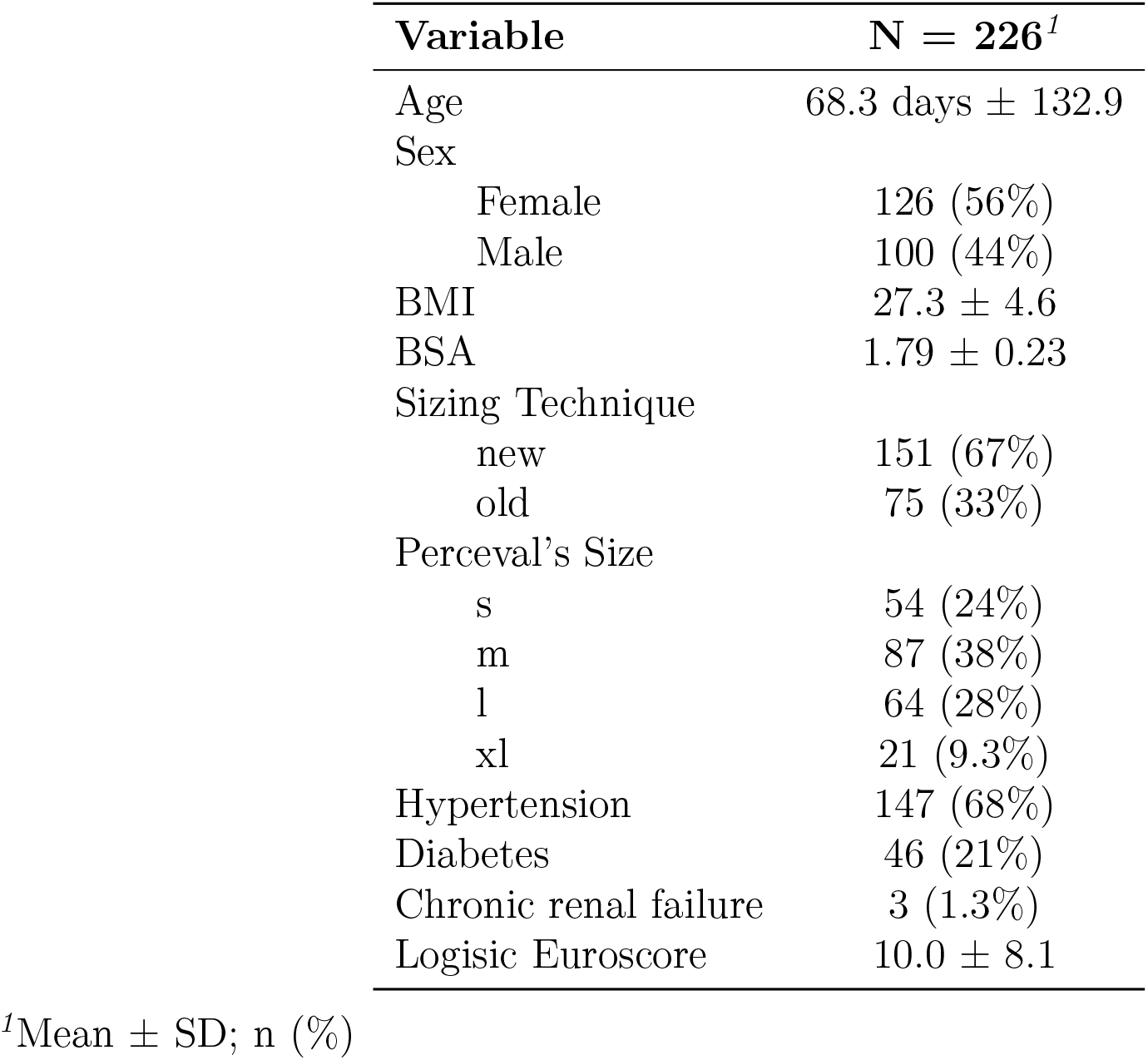
Patients’ Demografics And Baseline Charachteristics.

| Variable | N = 226<sup>1</sup> |
| --- | --- |
| Age | 68.3 days ± 132.9 |
| Sex |  |
| Female | 126 (56%) |
| Male | 100 (44%) |
| BMI | 27.3 ± 4.6 |
| BSA | 1.79 ± 0.23 |
| Sizing Technique |  |
| new | 151 (67%) |
| old | 75 (33%) |
| Perceval's Size |  |
| s | 54 (24%) |
| m | 87 (38%) |
| l | 64 (28%) |
| xl | 21 (9.3%) |
| Hypertension | 147 (68%) |
| Diabetes | 46 (21%) |
| Chronic renal failure | 3 (1.3%) |
| Logisic Euroscore | 10.0 ± 8.1 |

<sup>1</sup>Mean ± SD; n (%)

Over-sizing was calculated as:

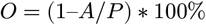

where *O* is percentage of over-sizing, *A* is a native virtual aortic ring surface area, *P* is the Perceval’s cross sectional area by manufacturer as published by our group before [6] (see Figure 1).

**Figure 1:**
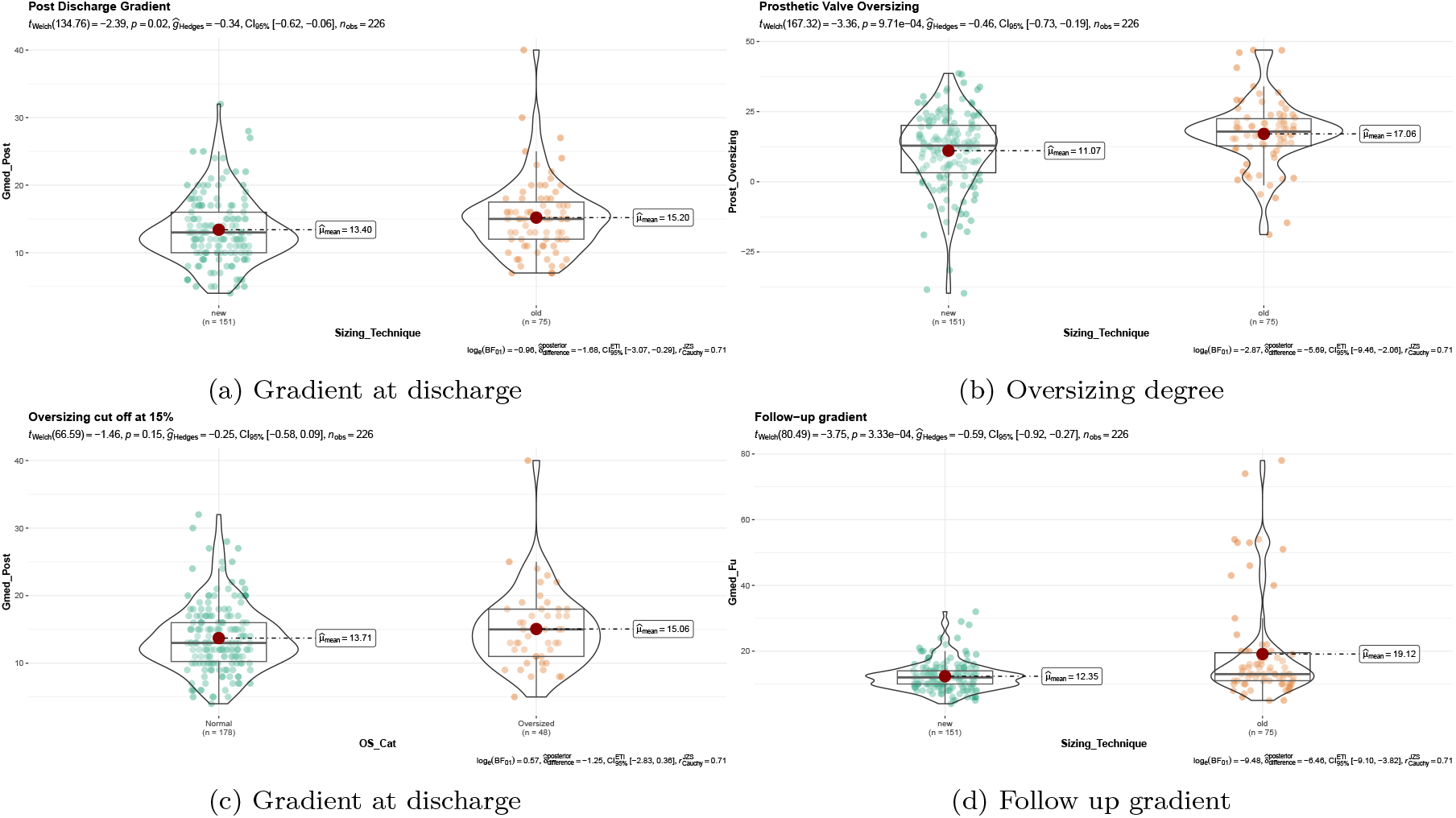
Prosthesis characteristics: a) gradient on prosthetic valve at discharge expressed in mmHg; b) calculated oversizing degree by sizing technique; c) discharge gradient diveded by cut off point (22.6%), d) follwo-up gradient by sizing technique.

**Figure 2:**
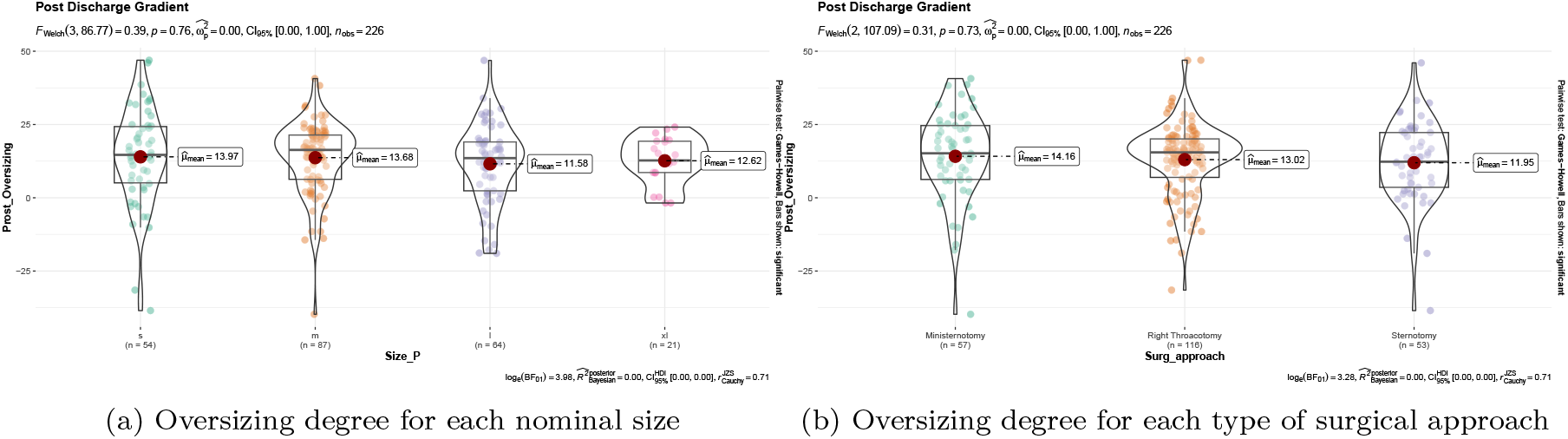
Prosthesis corelation to a) four sizes of Perceval prosthesis b) three surgical approaches.

### Surgical Procedure

The operations details are presented in Table 2. The aortic root was accessed through a high aortotomy at the level of the aortic fat pad or immediately below it [7]. The aortic valve leaflets were resected, and the annulus was de-calcified accurately (see supplementary Video 1).

**Table 2:** Surgical Caracheteristics.

| Variable<br>N = 151<sup>1</sup><br>N = 75<sup>1</sup> | new<br>old<br>p-value<sup>2</sup> |  |  |
| --- | --- | --- | --- |
| **Surgical Approach** |  |  | <0.001 |
| Ministernotomy | 53 (35 %) | 4 (5.3 %) |  |
| Minithoracotomy | 55 (36 %) | 61 (81 %) |  |
| Sternotomy | 43 (28 %) | 10 (13 %) |  |
| **Area** | 4.81 ± 1.06 | 4.79 ± 1.01 | >0.9 |
| **Perimeter** | 7.87 ± 0.88 | 7.96 ± 0.92 | 0.4 |
| **Prosthesis Oversizing** | 11 ± 14 | 17 ± 12 | 0.002 |
| **Perceval's Size** |  |  | <0.001 |
| s | 45 (30 %) | 9 (12 %) |  |
| m | 57 (38 %) | 30 (40 %) |  |
| l | 32 (21 %) | 32 (43 %) |  |
| xl | 17 (11 %) | 4 (5.3 %) |  |
| **CPB** | 106 ± 32 | 98 ± 35 | 0.003 |
| **Aortic X-Clamping** | 65 ± 19 | 62 ± 24 | 0.038 |
| **BSA** | 1.80 ± 0.24 | 1.77 ± 0.21 | 0.2 |
| **BMI** | 27.3 ± 4.4 | 27.1 ± 5.0 | 0.7 |

<sup>1</sup>n (% %); Mean ± SD<sup>162</sup> <sup>2</sup>Pearson's Chi-squared test; Wilcoxon rank sum test

### Surgical Sizing

Aortic annulus measurements approach were described by our group previously [8]. Briefly, old sizing was performed according to the instructions for use from manufacturer: the size of the prosthesis to be implanted was indicated by the sizer for which the transparent obturator passed through the annulus but the white obturator did not. New sizing was performed in our institution since 2017: the size of prosthesis to be implanted was indicated by the sizer for which the white obturetor passes with slight friction trough the native annulus. Thus, in practice it tends to under-size with respect to manufacturer recommendations of sizing [9].

### CT Protocol

For the purposes of this study, only patients with a suitable preoperative cardiac CT scan performed at our institution were considered. Cardiac CT was performed with a 320-slice multidetector CT scanner (Toshiba Aquilion ONE; Toshiba Medical Systems S.R.L., Rome, Italy). The scan was done using retrospective electrocardiogram gating in spiral technique. Contrast media (80 mL) was injected at 4 mL/s, and bolus tracking was used. The reconstructed slice width was 1 mm or less, and all of the measurements were done in diastole (75% of the cardiac cycle) to maximize the image quality and to avoid variation because the systolic images were not available for all patients. The patients’ CT scan Digital Imaging and Communications in Medicine (National Electrical Manufacturers Association, Rosslyn, VA) files were downloaded from our institutional images server, anonymized, and independently analyzed. The area and perimeter of the aortic valve VBR were measured on the three-dimensional multiplanar reconstructions as recommended [2] [10].

### Clinical and Hemodynamic Assessment

All patients were managed according to our routine institutional protocol. At the end of the surgical procedure, the prosthetic valve function was assessed by an expert cardiologist or cardiac anesthesiologist. Trans-thoracic echocardiography was always repeated before discharge and at at follow up. Continuous-wave Doppler was used to assess the flow velocity across the prosthetic valve. The clinical and hemodynamic outcomes are defined according to the Valve Academic Research Consortium 2 guidelines [11]. Procedure success was defined as having a single, normally functioning valve in the proper anatomical position. A mean transprosthetic gradient of 20 mm Hg or more was defined as an increased transprosthetic gradient. We have used standardized definition of structural valve degeneration for surgical and transcatheter bioprosthetic aortic valves [12]. Follow up is expressed in years.

### Statistical Analysis

Continuous variables are expressed as mean ± standard deviation, and categorical variables are expressed as percentages. The association of preoperative, intraoperative, and postoperative variables with an increased postoperative gradient was investigated by the Fisher exact test or Kruskal Wallis rank sum test (dichotomous variables) or by the unpaired Student t test (continuous variables). Normally distributed continuous variables were analyzed by the Mann-Whitney U test. Factors significantly associated to the end point of the study were included in a logistic multivariate regression model to ascertain their independent role. Also included were factors for which the univariate analysis gave a p value of 0.1 or less, or of known biologic significance, but failed to meet the critical level. Odds ratios (OR) and 95% confidence intervals (CIs) were calculated. Receiver operating characteristic (ROC) curves were calculated to single out the best cutoff value of prosthesis over-sizing predicting an increased postoperative gradient. The accuracy of the test was assessed by measuring the area under the ROC curve. The statistical significance of difference of the area under the ROC curve from that of the “line of no information” was evaluated by Mann-Whitney U statistic. A *p* value of less than 0.05 was considered significant. Statistical analysis was conducted using R Foundation for Statistical Computing with RStudio IDE.

## Results

### Procedural Outcome and Complications

Postoperative outcomes and complications are reported in Table 3. The operative mortality was 1 (0.4424779%). There were no prosthesis migration in neither of groups. There were no annular rupture or any kind of aortic root complication due to valve over-sizing. Postoperative PM implantation was documented in 12 (5.309735%) and it was not different between groups (p = 0.12). The mean follow up was lower in new group (3.3 ± 2.0 vs 9.7 ± 1.6 years, p < 0.01). At the discharge the patients who have used the new sizing technique had less gradient on the prosthetic valve (13.4 ± 5.0 vs 15.2 ± 5.5 mmHg, p = 0.02). The new sizing was less prone to degeneration which requires intervention at median follow-up (2 (1.324503%) vs 15 (20%), p < 0.01).

**Table 3:**
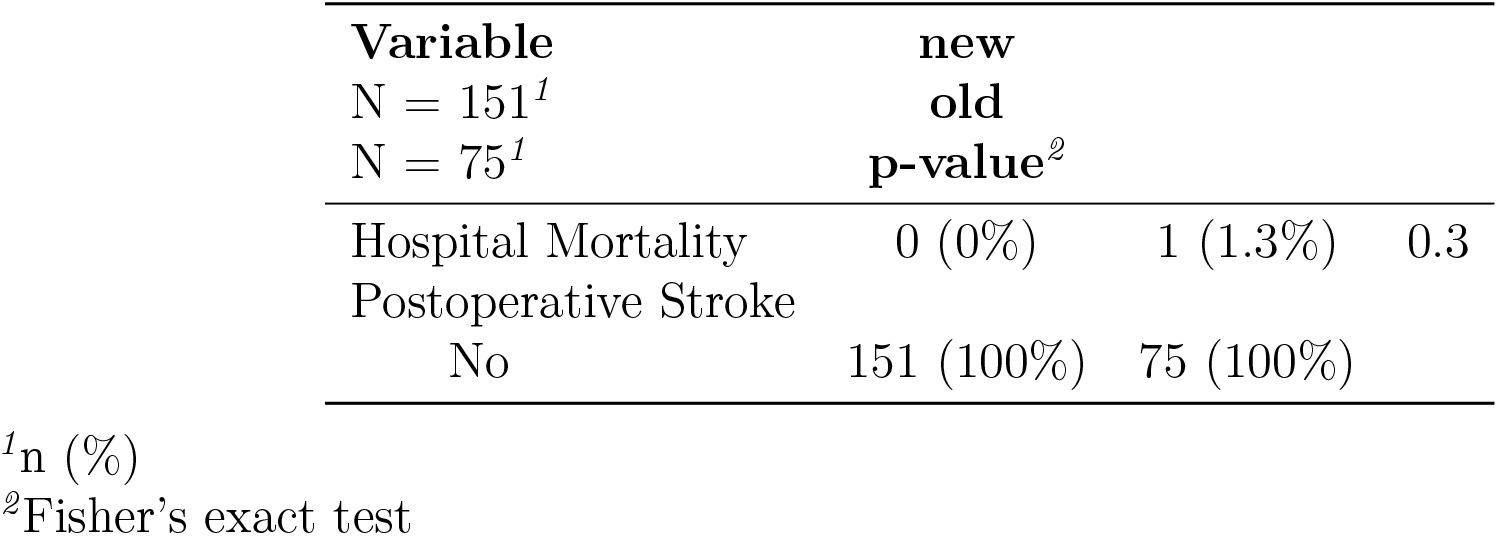
Postoperative Outcomes.

| Variable<br>N = 151<sup>1</sup><br>N = 75<sup>1</sup> | new<br>old<br>p-value<sup>2</sup> |  |  |
| --- | --- | --- | --- |
| Hospital Mortality | 0 (0%) | 1 (1.3%) | 0.3 |
| Postoperative Stroke |  |  |  |
| No | 151 (100%) | 75 (100%) |  |

<sup>1</sup>n (%)<sup>163</sup> <sup>2</sup>Fisher's exact test

### Prosthetic Valve performance

Gradient on prosthetic valve was significantly higher in the old group (p = 0.02). Over-sizing degree was significantly higher in the old group (p < 0.01). Gradient on prosthetic valve at 5.1 (95% CI: 0.4, 11.2) years of follow up was still significantly higher in the old group (p < 0.01, see Figure 1). Gradient on the valve at discharge and follow up were significantly correlating with over-sizing level (*p* < 0.05, see Figure 5, see Table 4).

**Table 4:**
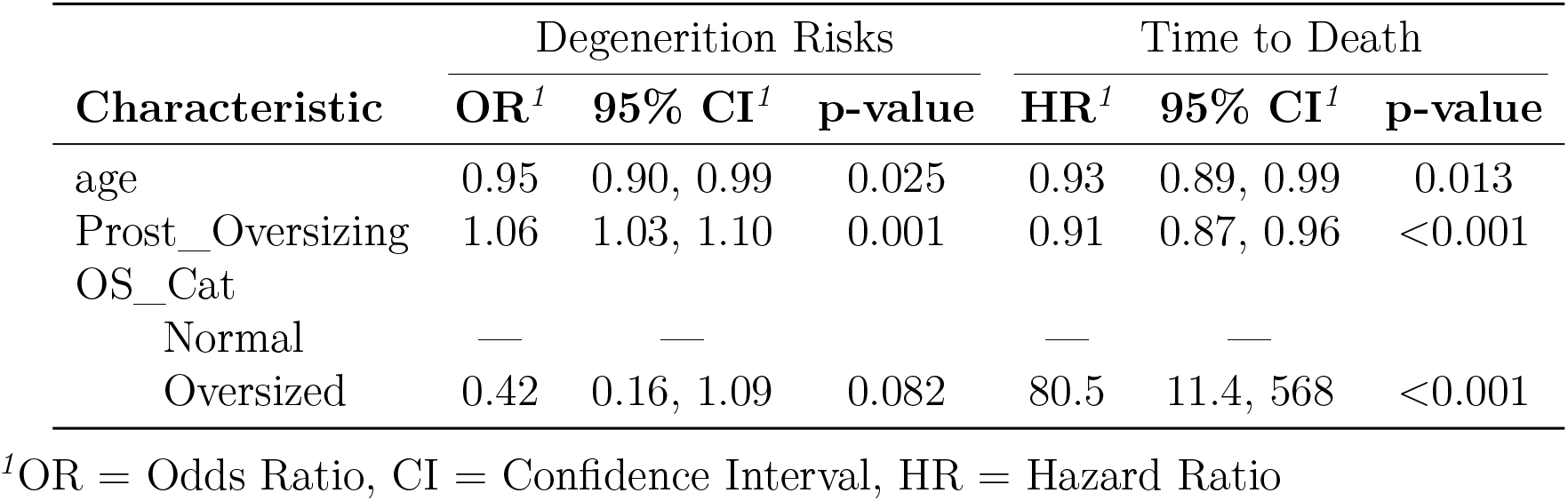
Risk Factors for Degeneration.

### Valve degeneration and survival analysis

At median follow up the survival probability was high (99.5575221 %, see Figure 4). At the available follow up there were no statistically different valve degeneration probability between two groups (p = 0.11, see Figure 4). However, when considering >22.6 % over sizing as a cut off (see Figure 3), there were significant degeneration difference being high in the old group (p < 0.01, see Figure 4).

**Figure 3:**
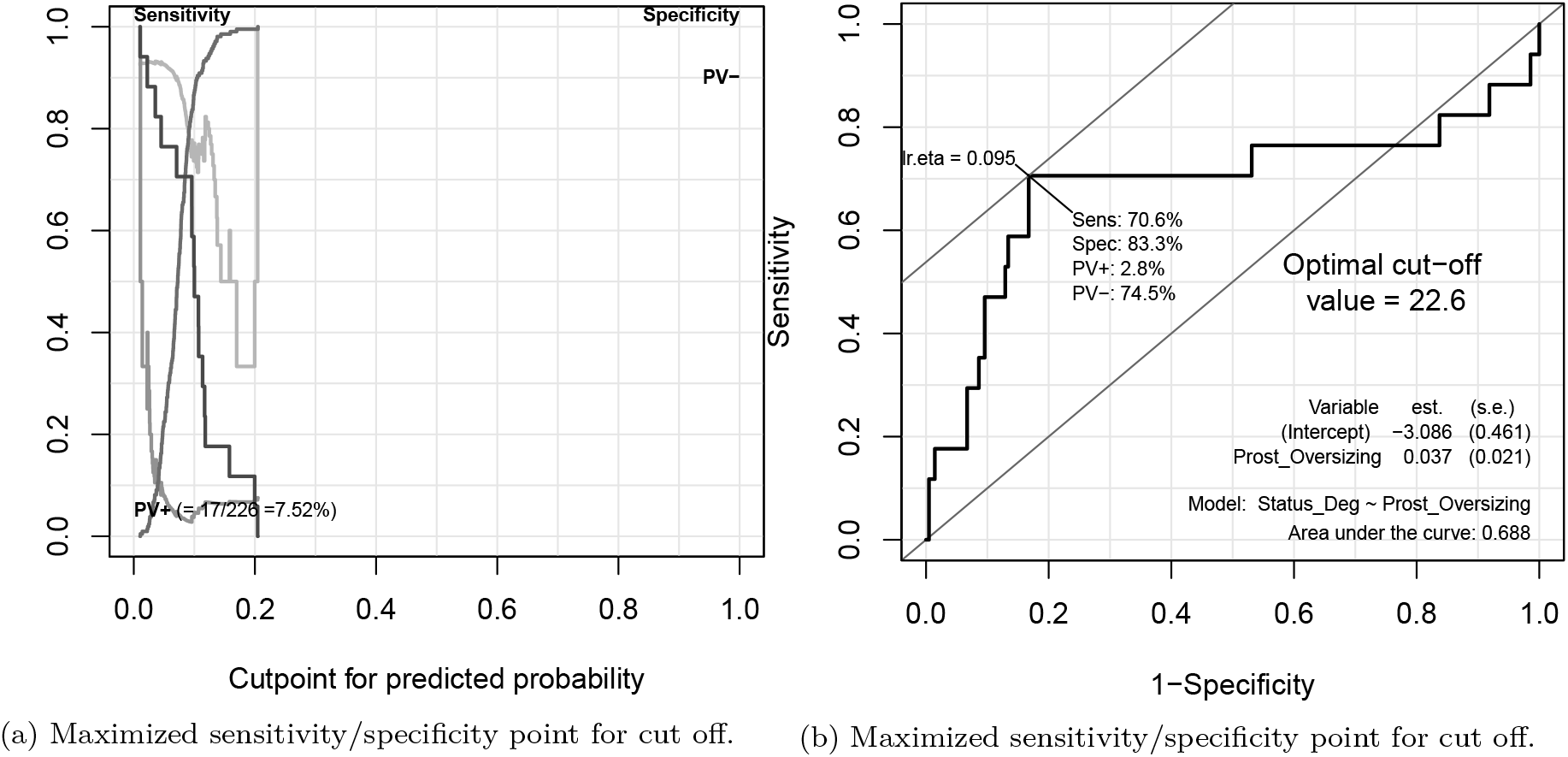
Over-sizing cut of analysis: a) cut-point of predicted probability; b) cut-off point and its parameters maximized (sensitivity, specificity) and model details that has been used to find the cut-off point.

**Figure 4:**
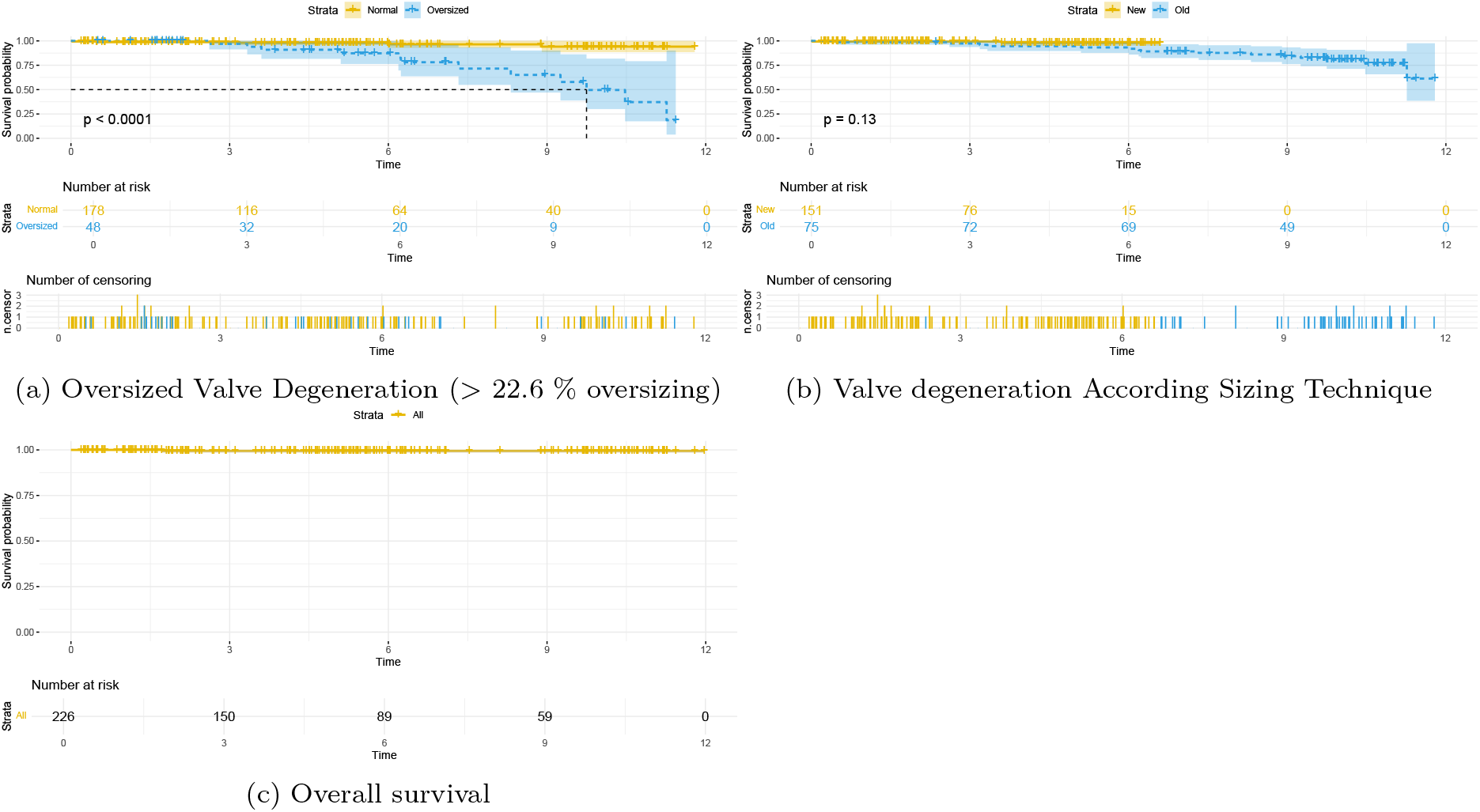
Kaplan Meier Curves: a) time to event of prosthesis structural valve degeneration(SVD) end/or surgical intervention; b)SVD devided by sizing technique; c) overall survival curve for cohort.

**Figure 5:**
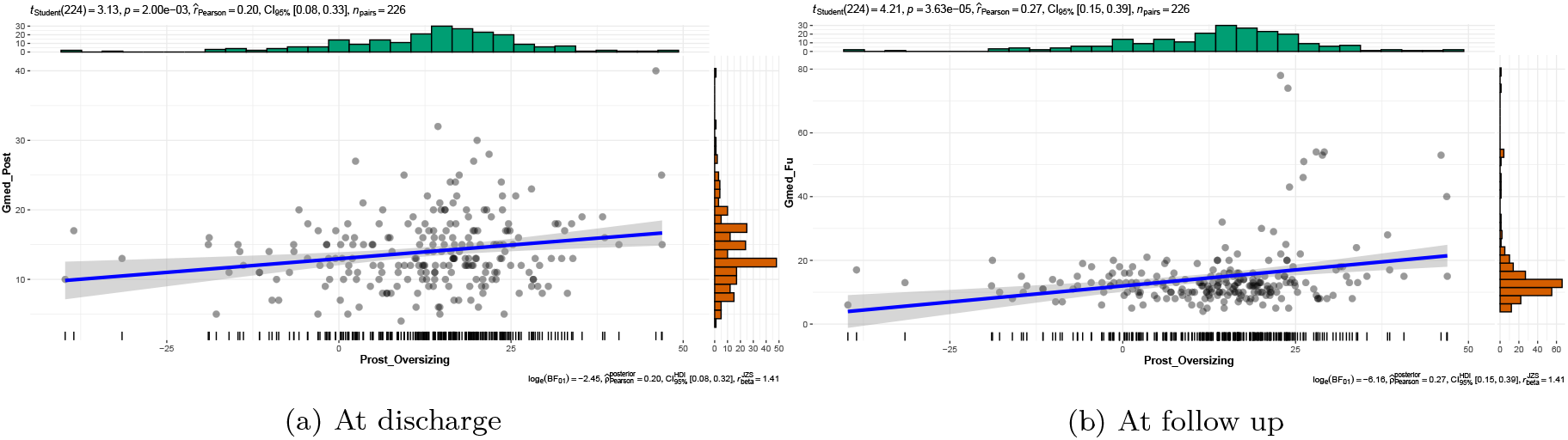
Oversizing and mean gradient correlation: a) at discharge; b) at follow-up.

## Discussion

Our study demonstrates that standard sizing of the Perceval valve is associated with increased transprosthetic gradients, higher degeneration probability if oversized.

Our study evidences the novel sizing technique is better alternative, which is safe and delivers less postoperative gradient, lower degeneration rate and long life cycle. It has been shown previously that over-sizing of 30% or more in the annular anatomical area was associated with a 16-fold increase of the risk of developing increased postoperative gradients [6]. In this study we found that even 22.6 % over-sizing is influencing negatively on valve degeneration (see Figure 4) and we recommended to avoid it. Intra-operative sizing for the Perceval valve, as recommended by the manufacturer instructions for use, does tend to over-size see Table 2. Surgical obturator-based sizing is a complex process that converts to a number several non-numeric inputs, including visual assessment, tactile feedback, stiffness of the cardiac tissues, the amount and distribution of calcium, the fragility of the aortic wall or the height of the coronary arteries and other relevant features based on operator. However, in our experience neither the size nor the approach do change the over-sizing degree (see Figure 2). Baert and coworkers [4] recently reported valve recoiling caused by excessive over-sizing in 4 patients (2.9% of their Perceval population). They suggested back in 2017 to modify the sizing process and to implant the valve size “Given by the sizer of which the white obturator pass the annulus with friction” [4]. We do use the novel sizing technique since 2017. In our opinion, CT guided sizing could be the best practice in order not to oversize more then 22.6 %. By given data, 15% over-sizing or less is well tolerated and has no accelerating influence on valve degeneration. Unlike previous studies our study focused on the early postoperative and follow-up periods. Increased gradients early after AVR can be related to many reversible factors, which disappear with time (unpublished data), and are often of limited clinical significance. However, increased gradients in patients with Perceval valves with known native annulus dimensions should not be neglected. In fact, as observed above, the presence of an increased gradient could indicate altered kinetics of the leaflets and may herald other negative outcomes, such as sub-clinical valve thrombosis and early degeneration, at least in some patients [13].

### Math explanation of oversizing

Area of a perfect circle is given by:

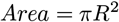

where R is the circle’s radius. Ellipse shape has an area give by this formula:

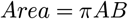

Where A and B are two radius-es of the ellipse. In order to achieve the same area ellipse should have shorter and longer radius’s multiplication equal to perfect circle’s radius square. On the other hand perimeter calculated as:

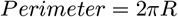

for perfect circle, and

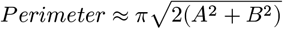

for elliptic shape. For every given prosthesis the perimeter is constant. Distorting is shape (other then perfect circle) yields to less area for given fix p erimeter, hence higher g radient. Oversize valve had external (heart’s) tissues radial forces which will distort it and constrain it into the non circular shape [13].

## Conclusions

New sizing technique is safe and reproducible. It seem to deliver better immediate and long term benefits for Perceval sutureless valve, less postoperative gradient, less probability or structural degeneration. Over-sizing above the 22.6 % should be avoided.

## Data Availability

All data produced in the present study are available upon reasonable request to the authors

## Acknowledgments

Conceptualization of the scientific question by RM, data were collected in mixed fashion by RM and GC. Manuscript drafting, editing and finalization by RM, GC and MS. Our study has some limitations: it is a single-institution retrospective analysis of prospectively collected data in a population of consecutive patients.

## Supplemental Material

## Notes

### Competing Interest Statement

The authors have declared no competing interest.

### Funding Statement

This study did not receive any funding

